# Psilocybin induces acute and persisting alterations in immune status and the stress response in healthy volunteers

**DOI:** 10.1101/2022.10.31.22281688

**Authors:** N.L. Mason, A. Szabo, K.P.C. Kuypers, P.A. Mallaroni, R. de la Torre Fornell, J.T. Reckweg, D.H.Y. Tse, N.R.P.W. Hutten, A. Feilding, J.G. Ramaekers

## Abstract

Patients characterized by stress-related disorders such as depression display elevated circulating concentrations of pro-inflammatory cytokines and a hyperactive HPA axis. Psychedelics are demonstrating promising results in treatment of such disorders, however the mechanisms of their therapeutic effects are still unknown. To date the evidence of acute and persisting effects of psychedelics on immune functioning, HPA axis activity in response to stress, and associated psychological outcomes is preliminary. To address this, we conducted a placebo-controlled, parallel group design comprising of 60 healthy participants who received either placebo (n=30) or 0.17 mg/kg psilocybin (n=30). Blood samples were taken to assess acute changes in immune status, and 7 days after drug administration. Seven days’ post-administration, participants in each treatment group were further subdivided: 15 underwent a stress induction protocol, and 15 underwent a control protocol. Ultra-high field magnetic resonance spectroscopy was used to assess whether acute changes in glutamate or glial activity were associated with changes in immune functioning. Finally, questionnaires assessed persisting self-report changes in mood and social behavior. Psilocybin immediately reduced concentrations of the pro-inflammatory cytokine tumor necrosis factor-α (TNF-α), while other inflammatory markers (interleukin (IL)-1α, IL-1β, IL-6, and C-reactive protein (CRP)) remained unchanged. Seven days later, TNF-α concentrations returned to baseline, while IL-6 and CRP concentrations were persistently reduced in the psilocybin group. Changes in the immune profile were related to acute neurometabolic activity as acute reductions in TNF-α were linked to lower concentrations of glutamate in the hippocampus. Additionally, the more of a reduction in IL-6 and CRP seven days after psilocybin, the more persisting positive mood and social effects participants reported. Regarding the stress response, after a psychosocial stressor, psilocybin blunted the cortisol response compared to placebo. Such acute and persisting changes may contribute to the psychological and therapeutic effects of psilocybin demonstrated in ongoing patient trials.

## Introduction

Substantial evidence has demonstrated that psychosocial stressors can activate the immune system, initiating inflammatory processes that may underlie certain psychiatric disorders[1-3]. An increasing number of studies have demonstrated that patients characterized by stress-related disorders such as depression, addiction, and post-traumatic stress disorder (PTSD) show elevated circulating concentrations of pro-inflammatory cytokines including interleukin (IL)-1α, IL-1β, IL-6, and tumor necrosis factor-α (TNF-α), as well as acute-phase proteins such as C-reactive protein (CRP)[2-8]. An inflammatory challenge (i.e. and endotoxin derived from *Escherichia coli*) results in the rise of of pro-inflammatory cytokines such as IL-6 and TNF-α in plasma and to induce depressive-like behaviors, including social withdrawal/disconnection and depressed mood in both healthy animals[9] and humans[10]. Further evidence suggests that heightened systemic inflammation may also contribute to non-responsiveness to current antidepressant therapies, resulting in treatment resistance[11, 12]. Overall, evidence supports that pharmacological treatments that decrease pro-inflammatory processes may hold therapeutic value in a wide range of neuropsychiatric diseases[13].

Serotonin (5-HT)2A agonist drugs, such as the classic psychedelics psilocybin, lysergic acid diethylamide (LSD), N,N-Dimethyltryptamine (DMT), ayahuasca (containing DMT), and 5-MeO-DMT, have been found to possess anti-inflammatory properties in preclinical models (for a review see Szabo [14]). In humans, one observational study found that inhalation of 5-methoxy-DMT (5-MeO-DMT) decreased IL-6 concentrations in saliva 1-1.5 hours after administration, with no effects on CRP or IL-1β[15]. The strongest evidence of psychedelics’ anti-inflammatory processes stems from one clinical study, which found a reduction of CRP, but not IL-6, in both depressed patients and controls, 48 hours after administration of ayahuasca compared to baseline[16]. In depressed patients, reductions in CRP correlated with a reduction in depressive symptoms. Accordingly, a growing number of clinical trials are finding promising results regarding psychedelics’ ability to treat a range of psychiatric disorders characterized by aberrant inflammatory profiles, such as (treatment-resistant) depression, addiction, and PTSD[17-19]. Taken together, evidence suggests that psychedelics possess systemic anti-inflammatory properties in humans that may contribute to their ability to alleviate (depressive) symptoms. However, the evidence of acute and persisting effects of psychedelic drugs on immune functioning, and their relationship with psychological outcomes such as increased mood and sociability is still preliminary.

If psychedelics reduce the symptomatology of stress-related disorders, and alter immune signaling, might they also modify an individual’s stress response? Indeed, it could be hypothesized that one way in which cytokine concentrations may be reduced is through lowered reactivity to psychosocial stressors. It has been established that psychosocial stressors increase peripheral cytokine concentrations, resulting in inflammation[2, 8]. Thus, a possible explanation for increased inflammation in the aforementioned disorders is a heightened stress response - and/or exposure to extreme stress - potentially in combination with a dysregulated hypothalamic-pituitary-adrenal (HPA) axis[20, 21]. A one or two-time ingestion of a psychedelic drug has been found to reduce symptomatology of stress-related disorders in clinical populations[22] and decrease feelings of stress in healthy populations [15, 23, 24]. Nevertheless, it has yet to be objectively and directly demonstrated whether an individual’s response to an acute psychosocial stressor is reduced in the days following ingestion of a psychedelic substance.

Finally, if psychedelics’ anti-inflammatory mechanisms give rise to symptom alleviation, it is of interest to understand the underlying biological mechanisms. It has been established that pro-inflammatory cytokines play a key role in immune system-to-brain signaling, influencing brain function, mood, and behavior[3, 25]. For example, peripheral cytokines have been found to cross the blood-brain barrier[3], influencing glutamate transmission[26] and inhibiting astrocyte glutamate uptake[27, 28], potentially leading to sustained glutamatergic activity, excitoxicity[29], and impaired synaptic plasticity in areas such as the medial prefrontal cortex and hippocampus. These brain areas are known to regulate stress and emotion[30, 31], and play a direct role in HPA axis functioning. These neuronal and glial alterations can give rise to large-scale neural network disruptions which may contribute to the pathophysiology of mood disorders[11, 32]. Interestingly, changes in glutamatergic transmission have been found to be altered by psychedelic drugs and suggested to play a role in the therapeutic efficacy of these substances in clinical trials[33-38]. However, whether such psychedelic-induced changes in glutamatergic or glial activity are associated with alterations in systemic inflammation has yet to be assessed.

The aim of the present double-blind, placebo-controlled, parallel-group design was fourfold: to assess the acute and persisting (7 day post) effects of the classic psychedelic psilocybin, on a range of inflammatory markers associated with the prognosis and therapeutic response of stress-related psychiatric disorders, including IL-1α, IL-1β, IL-6, IL-8, and TNF-α, as well as CRP, in healthy subjects. We further hypothesized that psilocybin-induced changes in immune status would be associated with psychosocial functioning. Acute glutamate and glial activity (concentrations of myo-inositol (mI)) were measured in the medial prefrontal cortex (mPFC) and hippocampus to assess whether acute changes in cytokine concentrations related with neurometabolic activity. Finally, to assess whether psilocybin reduces stress reactivity, 7-days following treatment administration participants underwent a stress-induction protocol (Maastricht acute stress test; MAST) or control protocol.

## Materials and Methods

A detailed description of the experimental procedure is provided in the Supplementary Methods and briefly summarized here and Figure 1.

**Figure 1.**
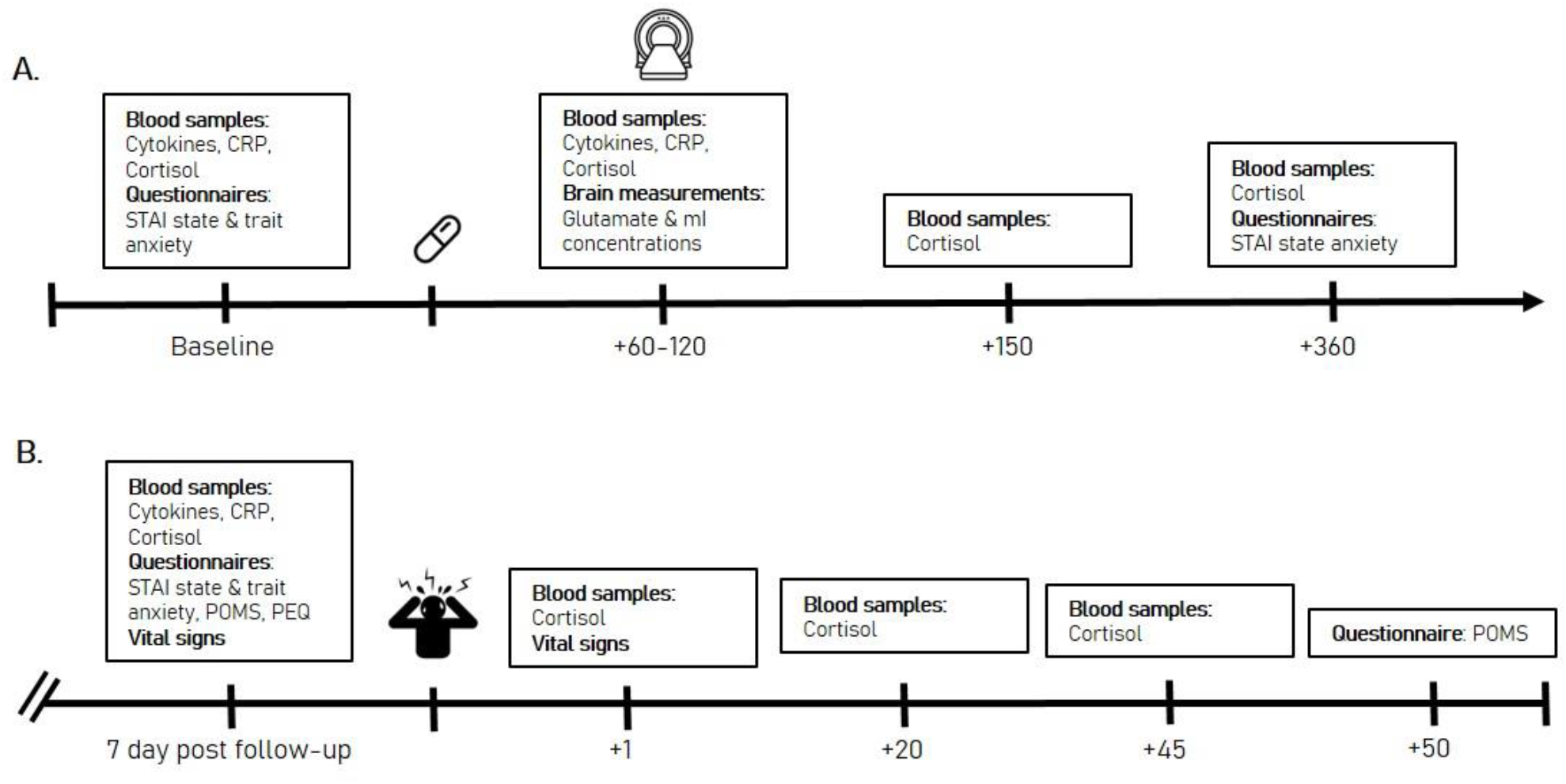
Experimental timeline. A) testing day 1, including psilocybin or placebo treatment. B) testing day 2, which took place 7 days after testing day 1. Timing is in minutes, relative to the treatment (psilocybin or placebo in A; stress induction or control protocol in B).

The study was conducted between July 2017 and June 2018 at Maastricht University, the Netherlands employing a balanced randomized (1:1), placebo-controlled, double-blind, parallel-group design. Sixty healthy participants, with previous experience with a psychedelic drug but not within the past 3 months, were allocated to a treatment condition (0.17 mg/kg psilocybin or placebo, p.o.), and then subsequently to a stress-induction condition (MAST or control). This resulted in 30 participants in the psilocybin condition, with 15 of those participants undergoing the MAST and 15 undergoing the control protocol, and 30 participants in the placebo condition, with 15 of those participants undergoing the MAST and 15 undergoing the control condition. All 4 groups were matched for age, sex, and education level. Full demographic information on the sample can be found in Table S1.

Participants visited the lab on three separate occasions. The first visit included a familiarization with testing day procedures. The second visit consisted of the formal testing day, with baseline blood samples (baseline), treatment administration (psilocybin or placebo), magnetic resonance spectroscopy (MRS) to assess glutamate and myo-inositol (mI) concentrations, and further blood samples (acute). The third testing day took place 7 days after the acute testing day, and included the final blood samples (follow-up), followed by the completion of the stress-induction protocol and physiological (cortisol, blood pressure) and psychological (subjective stress concentrations) stress response. Both the formal testing day and the follow-up testing day always started at 9:00am, to allow comparability of cortisol and inflammatory blood samples across testing days.

This study was conducted according to the code of ethics on human experimentation established by the declaration of Helsinki (1964) and amended in Fortaleza (Brazil, October 2013) and in accordance with the Medical Research Involving Human Subjects Act (WMO) and was approved by the Academic Hospital and University’s Medical Ethics committee. All participants were fully informed of all procedures, possible adverse reactions, legal rights and responsibilities, expected benefits, and their right to voluntary termination without consequences. The present data were part of a larger clinical trial (Netherlands Trial Register: NTR6505) of which parts have been previously published[33, 39].

### Immune assays

Venous blood samples were collected before treatment administration (baseline), after treatment administration on the acute testing day (during peak drug effects; 80 minutes), and 7 days after the acute testing day, to assess the concentrations of IL-1α, IL-1β, IL-6, IL-8, TNF-α, and CRP. Blood samples were collected in ethylenediaminetetraacetic acid (EDTA) tubes, centrifuged at 3500 rpm/min and the plasma fraction was isolated and stored at −20°C until assaying.

#### Cytokines

The simultaneous determination of IL-1α, IL-1β, IL-6, IL-8, and TNF-α plasma concentrations was performed using bead-based multiplexing technology using a XMAG-Luminex assay (Bio-Rad, Hercules, California, USA) (Bio-Plex Pro Human Cytokine Kit Panel). Briefly, standards, blanks, controls, and the participants’ samples were incubated with the suspension of beads covered with antibodies specific for the tested molecules. After the incubation and washing steps, the cocktail of biotinylated detection antibodies was applied, followed by incubation with streptavidin-phycoerythrin solution. The fluorescence signal was read on a BioPlex 200 equipment (Bio-Rad).

#### C-Reactive protein

CRP concentrations were assessed by a latex-enhanced immunoturbidimetric assay developed to accurately measure CRP concentrations in serum and plasma samples for conventional CRP ranges, using an ABX Pentra 400 Clinical Chemistry analyzer (ABX Horiba, Montpellier, France). Sample volumes were 4ul, analyzed in singles. The effective cut-off range for CRP level estimations was as follows: Reference range in adults (20-60 years) < 0.5 mg/dL; recommended cardiac risk assessment categories are low (<0.1 mg/dL), average (0.1 to 0.3 mg/dL), and high (>0.3 mg/dL).

### Stress Test

The MAST [40] was used at the 7-day follow-up day to induce acute stress. It is a reliable method to induce a strong autonomic, glucocorticoid and subjective stress responses[41]. The MAST combines physical stress induction, unpredictability, uncontrollability, and the social-evaluative nature of other stress induction protocols. In short, participants alternated between putting their hand in 2°C water for a period between 45 and 90 s and doing mental arithmetic (counting back from 2043 in steps of 17) while their faces were recorded and social-evaluative pressure (i.e. negative feedback) was provided by an experimenter unfamiliar to the participant. The control protocol was similar to the experimental protocol with the difference that water was lukewarm (36°C) and participants had to count from 1 to 25 at their own pace while no social pressure was applied. To determine individuals’ responses to the stressor, serum cortisol samples *(see neuroendocrine response)* and vital signs (heart rate (HR), systolic (SBP) and diastolic (DBP) blood pressure) were obtained before and following the MAST. Subjective ratings of anxiety were obtained before and after the MAST via the Profile of Moods Scale (POMS)).

### Neuroendocrine response

#### Acute

As a marker of HPA-axis activation and consequent immunomodulation[21], acute cortisol concentrations were assessed via venous blood samples, taken before treatment administration (baseline) and after treatment administration on the acute testing day (80, 150, and 360 minutes post).

#### In response to the stress test

Seven days post treatment, blood-cortisol was sampled again in response to the stress induction paradigm. This included a baseline sample 5 minutes before the MAST or control, and then samples 1, 20, and 45 minutes after the paradigm.

Blood samples were collected in ethylenediaminetetraacetic acid (EDTA) tubes, centrifuged at 3500 rpm/min and the plasma fraction was isolated and stored at −20°C until assaying. The reagent employed for plasma cortisol determination was Elecsys Cortisol II, Roche®. Samples were analyzed with a high-throughput immunochemistry module (Cobas® e801 Module, Roche) using electrochemiluminescence (ECL) technology.

### Subjective measures

#### State-Trait Anxiety Inventory (STAI)

The STAI[42] was completed by participants at baseline, at the end of the acute treatment day (360 minutes post-administration), and at follow-up. The STAI is a 40-item rating scale with a 4-point response format, ranging from 1 (*almost never*) to 4 (*almost always*) that is scored into two sub-scales (state anxiety and trait anxiety). For “state” anxiety questions, participants were asked to select the response for each item that best describes how they feel “right now, that is, at this moment”. For “trait” anxiety questions, participants were asked to select the response that best describes how they “generally feel, that is, most of the time”. Participants state anxiety levels were considered at all time points, but trait anxiety levels were only considered at baseline and follow-up.

The reverse-scored (e.g., “I feel pleasant”) and direct-scored (e.g., “I feel nervous and restless”) items are summed per each subscale to create a total score which ranges between 20 and 80, with higher scores indicating greater anxiety. Internal consistency coefficients for the scale have been shown to range from .86 to .95 and test-retest reliability coefficients from .65 to .75 over a 2-month interval, and considerable evidence attests to the construct and concurrent validity of the scale[42].

#### Profile of Mood States (POMS)

The POMS is a self-assessment mood questionnaire with 72 items, rated on a 5-point Likert scale, with 0 being ‘not at all’ to 4 ‘extremely’. Participants had to indicate to what extent these items were representative of their mood at that moment in time. Eight mood states are classified and quantified by calculating the sum score of associated items for each mood state, i.e., anxiety (9 items), depression (15 items), anger (12 items), vigor (8 items), fatigue (7 items), confusion (7 items), friendliness (8 items) and elation (6 items). For this study, only the anxiety subscale was considered.

#### Persisting Effects Questionnaire (PEQ)

The PEQ was completed by participants at the beginning of the follow-up day. It is a 143-item long scale aiming to assess changes in attitudes, moods, behavior, and spiritual experience which participants consider to be due to the experiences they had during the acute testing days [43]. Prior research has found that PEQ is sensitive to the prolonged effects of psychedelics, up to one year after ingestion[44]. The scale consists of six categories: *attitudes about life* (Number of items (N)= 26), *attitudes about self* (N= 22), *mood changes* (N= 18), *relationships* (N= 18), *behavioral changes* (N= 2), and *spiritual experience* (N= 59). Each category reflected positive and negative changes, resulting in 12 subscales. All items were rated on a 6-point scale (ranging from 0= *none* to 5= *extreme*). The scores of the resulting 12 scales (positive and negative scales for each of 5 categories) were assessed by calculating mean (SE) separately for each category[45].

### MRS acquisition and processing

Spectroscopic voxels were placed by a trained operator at the mPFC (voxel size = 25 × 20 × 17 mm3) and the right hippocampus (voxel size = 37 × 15 × 15 mm3). Spectra were acquired with the stimulated echo acquisition mode sequence (TE = 6.0 ms, TR = 5.0 s, 64 averages). Outcome measures for MRS were concentration ratios of glutamate and myo-inositol to total creatine (tCr, creatine + phospho-creatine).

Detailed information regarding image acquisition and MRS quantification is previously published[33]. Spectroscopy data was analyzed with LCModel version 6.3-1H.

### Statistical analyses

Statistical analysis was conducted in IBM SPSS Statistics 25 using a Linear Mixed Models analysis.

For the inflammatory response assessment and the STAI, the model included Session (baseline, acute, follow-up), Treatment (psilocybin or placebo), and Session × Treatment, with Treatment and Subject as fixed effects, Session as repeated effects, and a random intercept. A first order autoregressive covariance structure (AR1) was used.

If a significant main effect of Treatment or a significant Session × Treatment was observed, 2-sided independent samples *t* tests were used to compare means between treatments acutely (acute measurement – baseline measurement) and at follow-up (7 day measurement – baseline measurement). Due to violations of the assumption of normality (Shapiro wilks), concentration values of IL-6, CRP, and IL-8 were log transformed. One participant’s IL-6 values were excluded from analysis due to being an extreme outlier, which persisted even after log transformation (Table S2).

For the assessment of physiological and psychological responses to stress, the AR1 model included Timepoint (Cortisol: baseline, 1, 20, 24 minutes post stress test; Heart rate/blood pressure: baseline, post stress test; POMS anxiety ratings: baseline and 50 minutes post stress test), Treatment (psilocybin or placebo), and Stress condition (stress or no stress), Timepoint × Treatment × Stress condition, and Treatment × Stress condition, with Treatment and Subject as fixed effects, Timepoint as repeated effects, and a random intercept. If a significant main effect of Treatment or Stress condition, or a significant Timepoint × Treatment × Stress condition or Treatment × Stress condition was observed, independent samples *t* tests were used to compare means between stress conditions (cortisol: 1, 20, or 45 minute post stress test measurement – baseline measurement; heart rate/blood pressure and anxiety ratings: post stress test measurement-baseline).

Subscales on the PEQ were compared between the treatment groups using independent samples *t* tests.

Spearman’s correlations were conducted to assess the relationship between concentrations of neurometabolites and acute inflammatory markers which showed a treatment effect. To assess the potential relationship between changes in inflammatory markers and changes in mood and sociability, a canonical correlation was used, as this approach assesses the relationship between two multivariate data sets, allowing investigation of variables that may have multiple causes and effects, while also reducing the potential of type 1 error[46].

Statistical significance was set at a *P* value of less than .05.

## Results

### Demographic variables and relative glutamate/tCr and mI/tCr concentrations

Demographic information and relative glutamate/tCr and mI/tCr concentrations are all previously published elsewhere[33, 39] and briefly summarized here.

The psilocybin group (*n* = 30) and the placebo group (*n* = 30) did not differ in respect to demographic variables, such as sex, age, and drug use history (Table S1). Compared to placebo, relative glutamate/tCr concentrations were significantly higher in the mPFC (mean ± SE; psilocybin: 1.23 ± 0.02, *n* = 24; placebo: 1.14 ± 0.02, *n* = 28; *U* = 200.50, *p* = 0.01, *d* = 0.80) and significantly lower in the hippocampus (psilocybin: 0.77 ± 0.03, *n* = 21; placebo: 0.88 ± 0.03, *n* = 25; *U* = 163.50, *p* = 0.03, *d* = 0.69). There were no significant treatment differences when comparing concentrations of mI/tCr for the mPFC (mean ± SE; psilocybin: 0.78 ± 0.03, *n* = 25; placebo: 0.78 ± 0.03, *n* = 28; *U* = 346.50, *p* = 0.95, *d* = 0) or hippocampus (mean ± SE; psilocybin: 1.01 ± 0.05, *n* = 22; placebo: 1.04 ± 0.04, *n* = 24; *U* = 239.00, *p* = 0.58, *d* = 0.13).

### Psilocybin reduced inflammatory markers in a time and marker-dependent manner

#### TNF-α

Analysis revealed a significant Session × Treatment interaction on TNF-α (F_2, 78.47_= 3.06, *p* = 0.05). Compared to placebo, psilocybin significantly decreased TNF-α concentrations acutely (mean ± S.E.: psilocybin: −2.54± 0.80; placebo: 0.15 ±0.69; *t* = −2.54, df = 52, *p* = .014, *d* = 0.69) whereas concentrations 7 days’ post-treatment were unaffected (mean ± S.E.: psilocybin: −0.58 ± 0.86; placebo: .1.00 ±0.87; *t* = −1.27, df = 49, *p* = 0.20, *d* = 0.36) (Figure 2).

**Figure 2.**
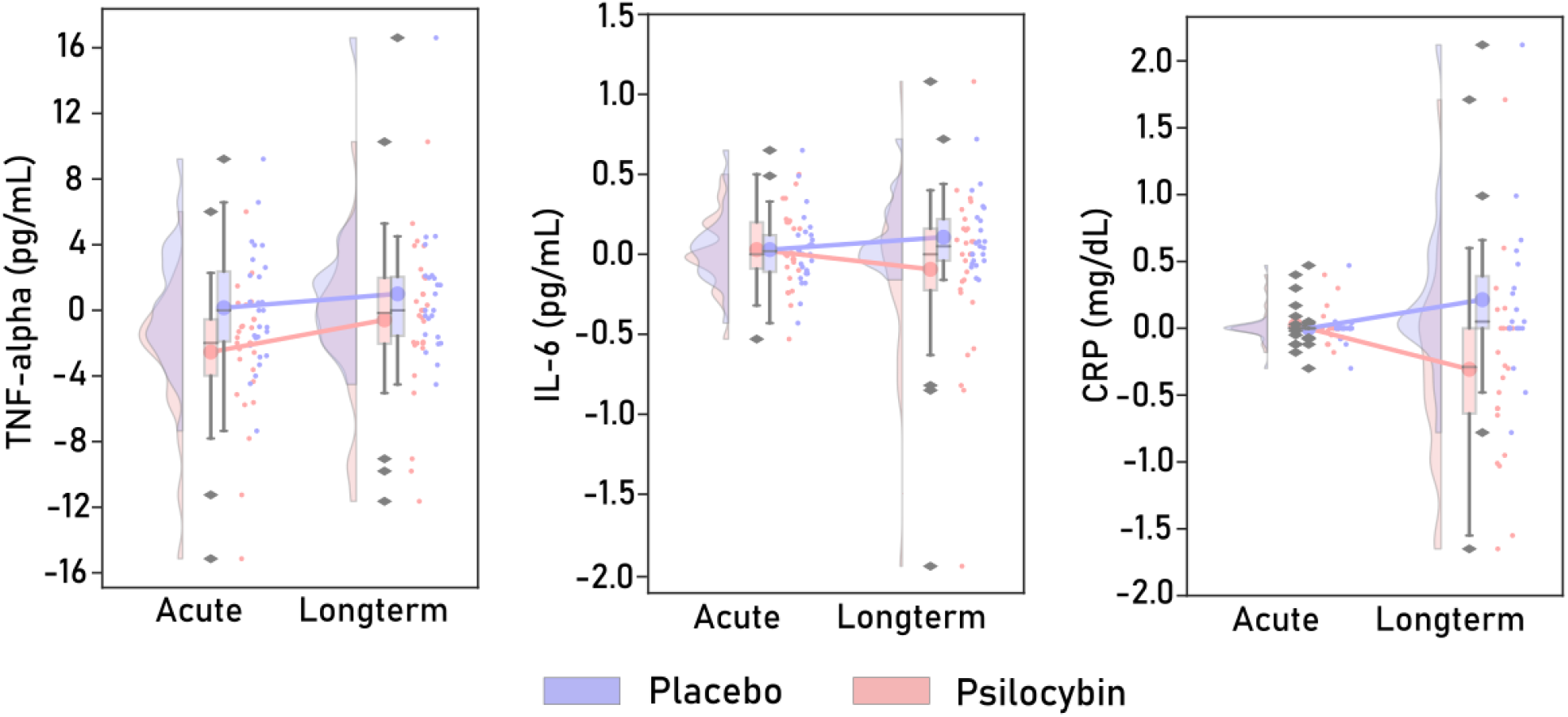
Raincloud plots displaying concentrations of immune markers (change from baseline) which demonstrated differences between treatment groups. The plot consists of a probability density plot, a boxplot, and raw data points. In the boxplot, the line dividing the box represents the median of the data, the ends represent the upper/lower quartiles, and the extreme lines represent the highest and lowest values excluding outliers. The code for raincloud plot visualization has been adapted from Allen, Poggiali [47]. Data points are change scores from baseline. CRP and IL-6 are log-transformed scores.

#### IL-6

Analysis revealed a significant Session × Treatment interaction on IL-6 (F_2, 97.59_= 3.21, *p* = 0.04). Compared to placebo, concentrations acutely did not differ after psilocybin (mean ± S.E.: psilocybin: 0.03 ± 0.05; placebo: 0.03 ±0.04; *t* = 0.02, df = 51, *p* = 0.98, *d* = 0), whereas psilocybin caused a trending decrease in IL-6 concentrations at follow-up (mean ± S.E.: psilocybin: −0.07 ± 0.54; placebo: .11 ± 0.04; *t* = −1.54, df = 35, *p* = 0.11, *d* = 0.44) (Figure 2).

#### CRP

Analysis revealed a significant Session × Treatment interaction on CRP (F_2, 85.53_= 6.52, *p* = 0.002). Compared to placebo, concentrations acutely did not differ after psilocybin (mean ± S.E.: psilocybin: 0.02 ± 0.03; placebo: 0.00 ± 0.03; *t* = 0.59, df = 43, *p* = 0.56, *d* =0.15), whereas psilocybin significantly decreased CRP concentrations at follow-up (mean ± S.E.: psilocybin: - 0.31 ± .15; placebo: 0.21 ± .14; *t* = −2.46, df = 39, *p* = 0.02, *d* = 0.77) (Figure 2).

#### IL-1β

There was no significant effect of Treatment on IL-1β (_F1,45.06_=3.70, p=0.06) or Session × Treatment interaction (F_2,66.58_=1.33, p=0.27). (Table S3).

#### IL-8

There was no significant effect of Treatment (F_1,55.24_=0.02, *p*=.87) or Session × Treatment interaction (F_2,95.47_=0.90, *p*=0.41) for IL-8. (Table S3).

### Psilocybin acutely activated the HPA axis, as indicated by increased cortisol concentrations up to 150 minutes post treatment administration

There was a significant Session × Treatment interaction on cortisol concentrations (F_3, 88.79_= 13.45, *p* = 0.000). Compared to placebo, psilocybin acutely increased concentrations from baseline, peaking at 80 minutes post administration (mean ± S.E.: psilocybin: 4.58 ± 1.02; placebo: −2.56 ± 0.87; *t* = 5.34, df = 53, *p* < 0.001, *d* = 1.43), and then began to fall at 150 minutes post administration (mean ± S.E.: psilocybin: 2.09 ± 1.13; placebo: −3.68 ± 0.97; *t* = 3.89, df = 52, *p* = < 0.001, *d* = 1.05). There were no differences between groups at 360 minutes post treatment (mean ± S.E.: psilocybin: −3.18 ± 0.97; placebo: −5.46 ± 1.06; *t* = 1.57, df = 52, *p* = 0.12, *d* = 0.43).

### Psilocybin blunted the neuroendocrine stress response to a psychosocial stressor 7 days post treatment administration

#### Cortisol

Analysis revealed a significant interaction between Treatment × Stress condition × Time on changes in cortisol (F_9,147.397_= 5.06 *p* < 0.001). Irrespective of psilocybin or placebo treatment, individuals in the stress condition had significantly higher increases in cortisol compared to the no-stress test (Table 1). That said, those in the psilocybin group demonstrated a lower cortisol increase than those in the placebo group (Figure 3).

**Table 1.** Cortisol values (ng/ml) after the stress paradigm, per treatment group and per stress condition. Values are change scores from baseline (taken 5 minutes prior to the stress test).

| Treatment | Minutes from stress test | Cortisol values (change from baseline) |  | *t* value | *p* | *d* |
| --- | --- | --- | --- | --- | --- | --- |
|  |  | Stress condition | No stress condition |  |  |  |
| Psilocybin | 1 | 0.96 (0.89) | -1.35 (0.44) | 2.32 | .032 | 0.85 |
|  | 20 | 0.23 (1.19) | -3.87 (0.63) | 3.09 | .005 | 1.14 |
|  | 45 | -0.83 (1.08) | -4.72 (0.69) | 3.08 | .005 | 1.14 |
| Placebo | 1 | 2.14 (0.55) | -1.54 (0.14) | 5.33 | <0.001 | 2.13 |
|  | 20 | 2.04 (1.17) | -2.51 (0.74) | 2.87 | 0.004 | 1.20 |
|  | 45 | 0.98 (1.22) | -3.37 (0.88) | 2.72 | .013 | 1.18 |

**Figure 3.**
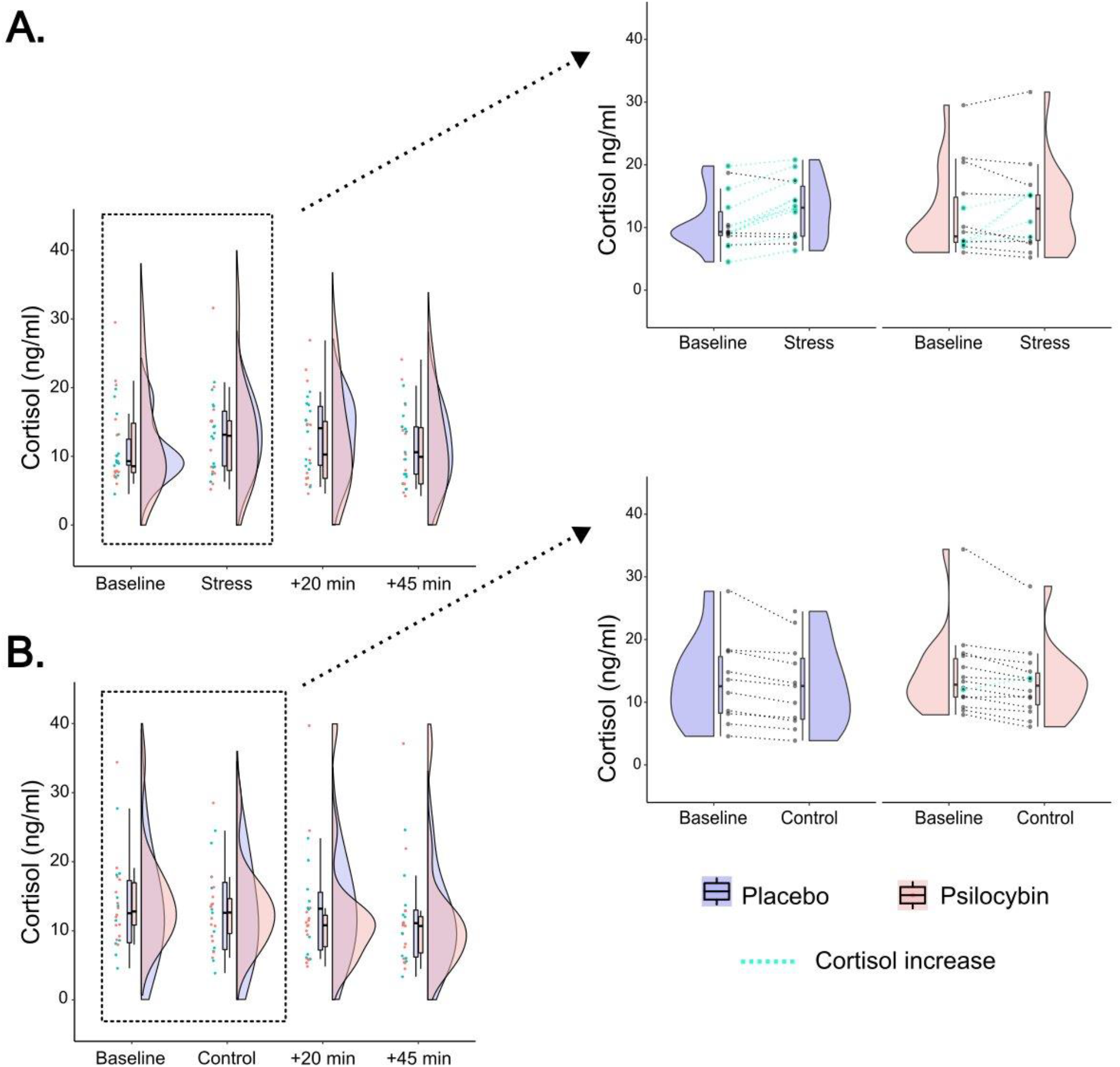
Neuroendocrine response (cortisol values) before, during, and after the stress (A) or the control (B) protocol, in those who received psilocybin or placebo. The left panel displays the cortisol response across all time points. The right panel zooms in, displaying cortisol concentrations before the stress/placebo protocol and during the stress/placebo protocol. The connecting lines demonstrate how individual participant’s cortisol concentrations changed over these two time points, and are separated by drug treatment condition (placebo or psilocybin). Blue lines indicate a cortisol increase.

#### Blood pressure and heart rate

Analysis revealed a significant interaction between Treatment × Stress condition × Time in diastolic blood pressure (F_3, 55.16_= 8.40, *p* = 0.000) and systolic blood pressure (F_3, 55.51_= 13.25, *p* = 0.000), but not beats per minute (F_4, 68.42_= 0.41, *p* = 0.80). In both the group that received the placebo treatment, and the group that received the psilocybin treatment, individuals in the stress condition had significantly higher increases in diastolic and systolic blood pressure (Table 2).

**Table 2.**
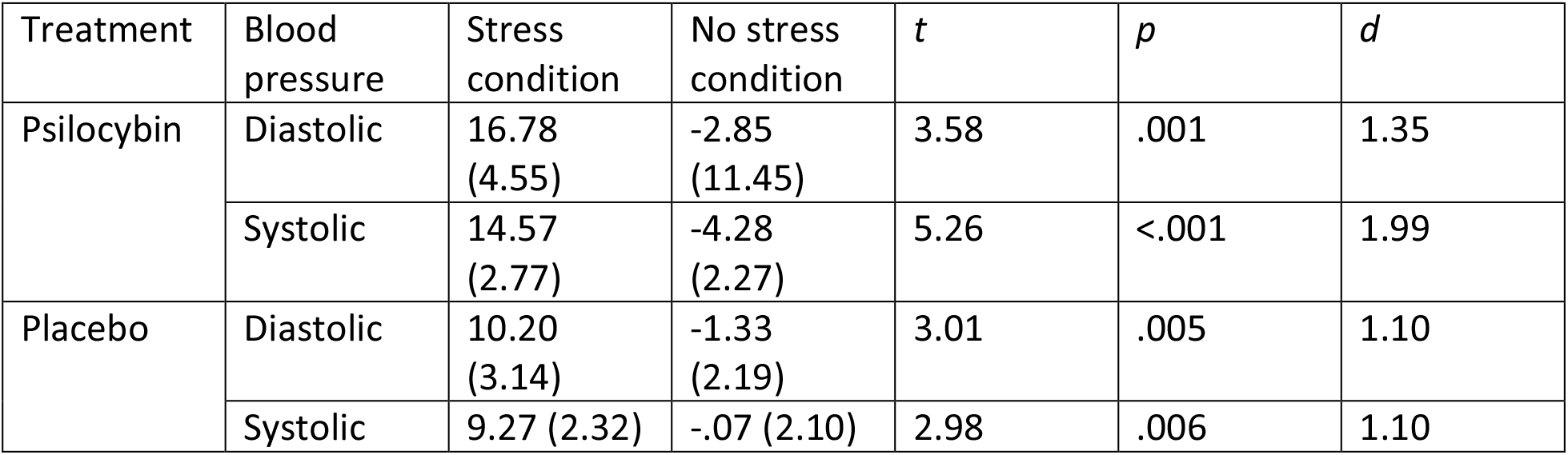
Blood pressure changes after the stress paradigm, per treatment group and per stress condition. Values are change scores from baseline (mean ± S.E).

**Table 3.** Canonical solution for inflammatory markers predicting persisting positive changes in mood and sociality for Functions 1 and 2. r_s_ > |0.45| and h^2^ > 45% are underlined and deemed valuable contributors.

| Variable | Function 1 |  |  |  |
| --- | --- | --- | --- | --- |
| | Coef. | $r_s$ | $r_s^2$ (%) | $h^2$(%) |
| Positive mood | .321 | <u>.962</u> | 92.54 | <u>92.54</u> |
| Altruistic/positive social effects | .6960 | <u>.992</u> | 94.41 | <u>94.41</u> |
| $R_c^2$ | | | 47.22 | |
| TNF- $\alpha$ acute change | .336 | .302 | 9.12 | 9.12 |
| IL-6 long-term change | -.402 | <u>-.855</u> | 73.10 | <u>73.10</u> |
| CRP long-term change | -.588 | <u>-.942</u> | 88.74 | <u>88.74</u> |

#### Rating of anxiety after the stress test (POMS)

Analysis revealed a trending interaction between Treatment × Stress condition × Time in ratings of anxiety 50 minutes after the stress test (F_3, 54.00_= 2.35, *p* = 0.08). Whereas individuals who had received the placebo treatment reported an increase in anxiety following the stress test compared to the control protocol (mean ± S.E.: stress: 1.14 (0.44); control: −0.71 (0.79); t=2.05, p=0.05, d=0.77), those who had received the psilocybin treatment did not show any difference in anxiety ratings when comparing responses following the stress test to the control procedure (mean ± S.E.: stress: - 0.51 (0.33); control: −0.27 (0.53); t=-0.42, p=0.67, d=0.15) Table S4.

### Psilocybin did not alter self-report ratings of state and trait anxiety, independent from the stress test

#### State

There was no significant effect of Treatment (F_1, 57.40_ = 1.23, *p* = 0.27) or Session × Treatment interaction (F_2, 78.87_ = 0.60, *p* = 0.55). Analysis did however reveal a significant Session effect on ratings of state anxiety (F_2, 78.87_ = 6.61, *p* = 0.002). When using paired samples *t*-tests to compare anxiety scores within-groups, it was found that compared to baseline, participants in both treatment groups reported significant reductions in state anxiety at the end of the acute testing day (psilocybin: *t* = 3.07, df = 29, *p* = 0.005, *d* = 0.52; placebo: *t* = 2.22, df = 28, *p* = 0.03, *d* = 0.34), whereas state anxiety levels at the 7 day follow-up did not differ compared to baseline (psilocybin: *t* = 1.31, df = 29, *p* = 0.20, *d* = 0.29; placebo: *t* = 1.15, df = 27, *p* = 0.26, *d* = 0.19).

#### Trait

There was no significant effect of Treatment (F_1,57.12_=0.06, *p*=0.81) or Session × Treatment interaction (F_1,56.26_=0.37, *p*=0.55).

### Persisting effects of psilocybin on changes in attitudes, mood, sociability, behavior, and spiritual experience

Those who received psilocybin had significantly higher ratings of positive changes on attitudes about life (*t*(57)=4.42, *p*<0.001, *d*=1.15), positive attitudes about the self (*t*(57)=4.45, *p*<0.001. *d*=1.08), positive mood changes (*t*(57)=3.90, *p*<0.001, *d*=1.02), altruistic/positive social effects (*t*(57)=3.80, *p*<0.001, *d*=0.98), positive behavior changes (*t*(57)=5.70, *p*<0.001, *d*=1.22), and increasing spirituality (*t*(57))=3.12, *p*=0.003, *d*=0.81) 7 days after psilocybin, compared to the placebo group.

There were no significant differences in changes in negative attitudes about life, negative attitudes about the self, negative mood changes, antisocial/negative social effects, negative behavior changes, and decreasing spirituality. For mean and statistical values see Table S5.

## Correlations

### Psilocybin-induced changes in TNF-α correlate with relative concentrations of glutamate/tCr in the hippocampus

A significant positive correlation was found between acute changes in TNF-α, and concentrations of glutamate/tCr in the hippocampus (R=.514, *p*=0.006, n=27), indicating that after psilocybin, the more of a reduction of TNF-α, the less hippocampal glutamate modulation occurs (Figure 4). There were no significant correlations between changes in IL-6 and glutamate/tCr concentrations in the mPFC and hippocampus after psilocybin (all *p*>0.15).

**Figure 4.**
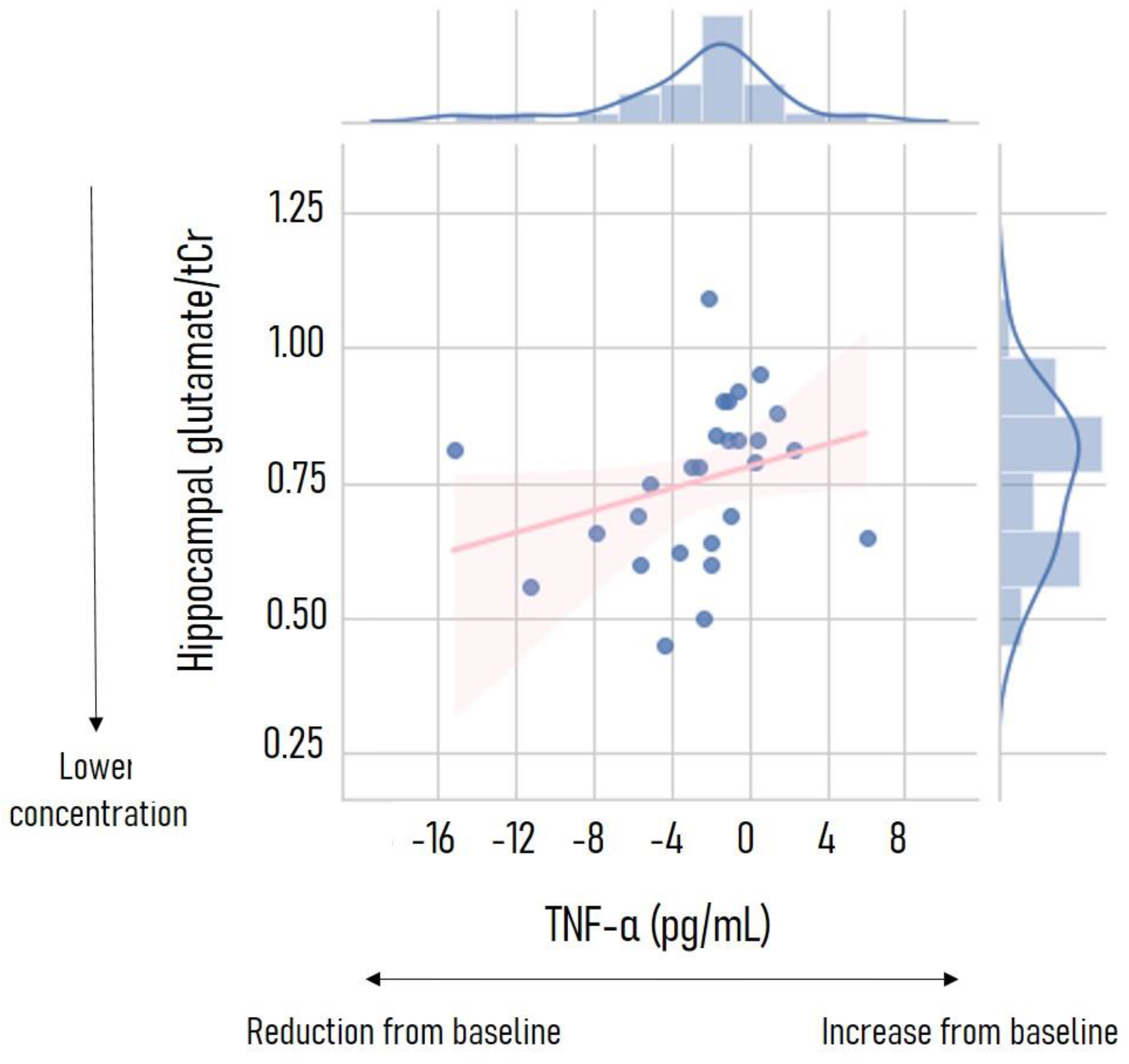
Scatter plot depicting relationship between acute changes in TNF-α (pg/mL) and hippocampal glutamate/tCr concentrations, in the psilocybin condition.

### Psilocybin-induced changes in IL-6 and CRP predict persisting positive changes in mood and sociality

A canonical correlation analysis was conducted using the three markers of immune functioning (change scores from baseline) as predictors of reported persisting changes in mood and social relationships.

The analysis yielded two functions with squared canonical correlations (*R*_c_^2^) of .472 and .115 for each successive function. The full model across all functions was statistically significant (*F*_6, 32.00_= 2.47, *p*=.04), explaining 54% of the variance. From this model, the first function was considered noteworthy, explaining 47.22% of the variance. The dominant contributors to the model were both persisting positive changes in mood and social relationships, whereas the dominant predictors were persisting decreases in IL-6 and CRP.

## Discussion

The present study demonstrates that a single, moderate dose of psilocybin has both acute and persisting effects on the immune profile and the stress response in healthy volunteers. Regarding the immune profile, psilocybin immediately reduced concentrations of the pro-inflammatory cytokine TNF-α. This effect was specific, as the other assessed inflammatory marker concentrations remained unchanged. Seven days later, TNF-α concentrations returned to baseline while IL-6 and CRP concentrations were reduced after psilocybin, compared to placebo. When investigating whether such changes in the immune profile were related to acute neurometabolic activity, it was found that acute reductions in TNF-α were linked to lower glutamate/tCr concentrations in the hippocampus, but not to glial activation (mI), suggesting that changes in plasma TNF-α concentrations may be attributed to peripheral immune modulation. When assessing whether changes in the inflammatory response related to persisting changes in psychological outcomes, it was found that the more of a reduction in IL-6 and CRP seven days after psilocybin, the more persisting positive mood and positive social effects participants reported. Regarding the stress response, after a psychosocial stressor, participants who had previously received psilocybin demonstrated a blunted cortisol response, and reported marginally lower concentrations of anxiety, compared to those who had received the placebo treatment.

### Acute and persisting effects of psilocybin on the immune profile

The effect of psilocybin on the immune profile of participants was two-fold: 1) acute changes were characterized by significantly decreased plasma concentrations of TNF-α, whereas 2) long-term changes were associated with decreased concentrations of circulating IL-6 and CRP. These changes are consistent with previous *in vitro* and animal *in vivo* studies demonstrating the anti-inflammatory properties of serotonergic psychedelics[14, 17, 29, 48-50]. TNF-α is an early response cytokine and a crucial, integrated part of inflammatory innate immune responses. Its modulation is therefore rapid and falls under tight control[51, 52]. As the acute, down-regulatory effect of psilocybin on circulating TNF-α was associated with increased concentrations of plasma cortisol (see supplementary results), one possible interpretation is the cortisol-induced modulation of this cytokine. Cortisol is known to be quickly and robustly induced by both psychological and physical stress and has a crucial role in stress adaptation and the restoration of homeostasis following acute stressors[53]. Acute, peak doses of psilocybin[54], and its serotonergic psychedelic analogs are known to induce cortisol release in humans[15], and thus may have a stress-mimicking effect via HPA-axis modulation[55].

Furthermore, cortisol and related glucocorticoids can rapidly and effectively suppress inflammation and have been used in the clinical therapy of both acute and chronic inflammatory diseases[56]. The observed, prompt decrease in plasma TNF-α might be the direct result of the cortisol peak elicited by psilocybin administration. Another possible interpretation is that psilocybin exerts its anti-inflammatory effects via the 5-HT2A and/or sigma-1 receptors of circulating immune cells and thereby causes rapid changes in blood TNF-α concentrations. Previous preclinical studies with similar psychedelic analogs support this hypothesis [15, 48-50, 57].

IL-6 is a pleiotropic cytokine that has a dominantly pro-inflammatory effect in systemic and tissue-specific immune regulation[58]. IL-6 is a direct inducer and regulator of CRP in the liver, and thus blood IL-6 and CRP concentrations are strongly correlated (as also demonstrated in our data, see supplementary results), and CRP is considered an important circulating biomarker of inflammation in clinical practice[59]. Since cortisol has a short biological half-life, and systemic cortisol-mediated effects normally last only for a couple of hours in humans, long-term decreases in the concentrations of both IL-6 and CRP are unlikely to be mediated by cortisol. Long-term immunomodulation by psilocybin via the 5-HT2A and/or sigma-1 receptors of immune cells offers a plausible explanation for the observed phenomenon and is also consistent with previous reports on the anti-inflammatory capacity of psilocybin and related psychedelic tryptamines, as discussed above (reviewed in[14, 17]). Finally, the persisting anti-inflammatory effects may also be the result of neuro-immunoregulatory feedback loops through HPA-axis modulation that involve cortisol in the acute phase and possibly other neuroendocrine regulators as well as cytokines in the long-run[53, 60].

IL-6 and CRP are established, robust markers in major depressive (MDD) and anxiety disorders where elevated concentrations are associated with more severe symptoms and symptom-dynamics[61-66]. Thus, the long-term effect of psilocybin on circulating biomarkers of inflammation may reflect an important biological component of its documented, persistent antidepressant effects[67, 68], and suggests a similar mechanism for other tryptamines[69, 70]. Relatedly, compared to those who received placebo, 7 days after psilocybin administration, participants reported a range of persisting positive effects which they attributed to psilocybin administration. A relationship was found between persisting reductions in CRP and IL-6, and increases in self-rated positive mood and social effects, two domains which are particularly interesting in relation to immune functioning. Namely, previous experimental work has demonstrated that when healthy participants undergo an inflammatory challenge, they show increases in depressed mood, and report feeling more socially disconnected from others[10, 71, 72]. Such findings are in line with research which suggests that pro-inflammatory cytokines initiate a “sickness behavior”, characterized by social withdrawal, fatigue, anhedonia, and reduced appetite, which is reversible once cytokine concentrations are reduced[73, 74]. Importantly, this cytokine-induced sickness behavior is an innate, adaptive immune system response, thought to facilitate recuperation from illness by conserving energy to combat acute inflammation[75]. That said, this behavior can also become pathological, for example if it is exaggerated in intensity (cytokines produced in high quantities) or duration (cytokines produced for a longer duration than normal)[73]. Accordingly, there is growing evidence that disorders such as depression are associated with significant elevations in circulating concentrations of pro-inflammatory cytokines, especially IL-6, as well as CRP[61, 76, 77]. As such, cytokine modulators have been suggested to be novel drugs for depression[78, 79], particularly if patients’ baseline inflammatory concentrations are high[80]. In trying to understand the relationship between inflammation and depressive symptomology, previous work has found that inflammatory-induced feelings of social withdrawal mediate increases in depressive mood during an inflammatory challenge[71]. Interestingly, it has been repeatedly found that a single administration of a psychedelic drug, like psilocybin, lysergic acid diethylamide (LSD), ayahuasca, or the mixed 5HT_2A_ agonist/ monoaminergic releaser 3,4-methylenedioxymethamphetamine (MDMA) induces prosocial effects and increases in feelings of connectedness in both healthy participants (for a review see Preller and Vollenweider [81]) and patients[82]. These psychedelic-induced prosocial effects have been repeatedly found to relate to persisting (therapeutic) outcomes[82-84]. Finally, the only other study to-date which assessed the relationship between inflammatory markers and persisting psychological effects after psychedelic administration, found that the larger the reduction of CRP, the lower the ratings of depression 48 hours after ayahuasca treatment[16]. Thus taken together, evidence suggests a relationship between immune system functioning, prosocial behavior, and psychological well-being after psychedelic intake, which could account for some of the immediate and persisting therapeutic responses seen in psychedelic-assisted clinical trials.

### Relationship between psilocybin-induced changes in the inflammatory response, and brain neurometabolites

Our findings about the acute and persistent effects of psilocybin administration on circulating cytokines show both an early-immediate (TNF-α) as well as long-term systemic, anti-inflammatory modulation (IL-6 and CRP). In order to be able to contextualize these results in a psycho-neuroimmunological frame of interpretation, we assessed the concentrations of mI and glutamate in several brain areas implicated in stress and emotion regulation, and correlated those which demonstrated a treatment effect the with the immune biomarker results.

Myo-inositol is an important marker of glial activation reflecting the inflammatory activity of the immunocompetent cells of the brain: astrocytes and microglia (reactive gliosis)[85, 86]. As mI/tCr concentrations did not display a treatment effect, it is unlikely that glial activation in the mPFC and hippocampus contribute to the psilocybin-altered immune response. Contrarily, psilocybin induced region-dependent alterations in glutamate, thus we next correlated glutamate/tCr concentrations with acute (TNF-α) concentrations in relevant areas of the cortex. We found a positive relation between acute TNF-α and glutamate/tCr concentrations in the hippocampus. Cortical glutamate concentrations and glial activity (especially that of astrocytes) are tightly connected, and inflammation has been shown to influence glutamate neurotransmission in the brain[27]. Furthermore, it has also been reported that inflammation and dysregulated glial activation can lead to increased release and abnormal clearance of glutamate in the synaptic space contributing to the pathophysiology of mood disorders, such as MDD[27]. According to previous reports, brain TNF-α and glutamate act synergistically and their concentrations are correlated, a phenomenon that has been suggested to be a core mechanism in inflammation-related neurodegenerative processes[87], thus our findings are complementary by demonstrating that after psilocybin administration, greater reductions of TNF-α were associated with less hippocampal glutamate modulation. We thus speculate that peripheral psilocybin administration likely does not affect the glial TNF-α-glutamate axis and does not induce pathological/inflammatory glia modulation.

In sum, our findings show some degree of area-specificity of acute psilocybin effects that are mostly related to the hippocampus via TNF-α and glutamate/tCr. The observed effects are not persistent and do not indicate a global, all-encompassing feature with regards to glial cell activation. This might be the consequence of a direct, local effect of psilocybin (in the hippocampal area), or on the other hand, may be an issue of statistical power due to sample numbers. As such, further investigations into the potential cortical glia-modulatory effects of psilocybin is highly warranted.

### Persisting effects of psilocybin on the stress response

Compared to the no-stress control protocol, the MAST induced significant autonomic responses including increases in systolic and diastolic blood pressure, and increases in cortisol, in both those which received psilocybin, and those that had received placebo. Such findings are in line with previous studies utilizing the MAST in a healthy population[40, 88-92]. In contrast to the previous studies whom also demonstrated a strong increase in subjective concentrations of state anxiety after the MAST, ratings of anxiety did not reach significance when comparing the stress to no stress condition. That said, in the current study anxiety ratings were taken 50 minutes after the stress test, a timeframe in which previous studies have demonstrated participants ratings of anxiety have returned to baseline[88].

When comparing autonomic responses on the MAST between the treatment groups, those who had received psilocybin instead of placebo demonstrated a blunted cortisol response (T_1_: *d* = 0.85 vs *d*=2.13, respectively), whereas they demonstrated a marginally higher cardiovascular reactivity (systolic blood pressure: *d*=1.35 vs *d*=1.10; diastolic blood pressure: *d*=1.99 vs *d*=1.10), and marginally lower ratings of anxiety (*d*=0.15 vs *d* =0.77). The specificity of these findings are interesting to hypothesize on. Namely, the MAST was developed to combine both physical and psychological stress components, which activate both the sympatho-adrenal-medullary (SAM) and the HPA axis[40]. The SAM is considered the “fast response” to a stressor, acting via direct and fast sympathetic nerve stimulation, which results in secretion of catecholamines such as adrenaline and noradrenaline, and increases in heart rate, blood pressure, and respiration frequency. This quick response helps the body focus its resources, giving rise to the well-known “fight or flight” response[93]. The second stress response pathway is slower, acting via the HPA axis which commences with the perception of stress[94], resulting in the hypothalamus releasing corticotropin releasing hormone, which triggers excretion of adrenocorticotropic hormones by the pituitary gland, and ultimately promotes release of cortisol in the bloodstream via the adrenal glands[95]. As such, when assessing whether psilocybin alters the acute stress response, our data could suggest a stress response-dependent effect. Namely, in the days following psilocybin administration, when exposed to a stressor participants display a normal (or potentially marginally heightened) SAM response, demonstrated via comparable cardiovascular reactivity, and a blunted HPA axis response, demonstrated by lower cortisol secretion.

Hyperactivity of the HPA axis, and resultant hypercortisolism, are one of the most consistent biological findings in regards to stress-related disorders[96-98]. The path from hyperactive HPA axis to pathology is suspected to be due to sustained concentrations of cortisol, which reduce brain derived neurotrophic factor (BDNF), ultimately proving toxic to neurons in key brain regions which control the HPA axis such as the hippocampus[99]. Such damage to the hippocampus may result in an inability of the region to exert inhibitory control of the HPA axis, leading to a further increase in circulating cortisol, and more hippocampal damage[98]. In light of the current investigations of psychedelics for stress-related disorders, it is interesting to speculate on whether the findings of the current study may highlight a potential therapeutic mechanism of these drugs, i.e., downregulation of the HPA axis. Multiple studies have demonstrated 5-HT_1A_ and 5-HT_2A/2C_ agonists, including psychedelic drugs[100, 101], activate the HPA axis, stimulating ACTH and cortisol release, and interacting with receptors in control of glucocorticoid secretion-namely glucocorticoid (GR) and mineralocorticoid (MR) receptors [102-105]. Additionally it has been demonstrated that both the 5-HT_1A_ receptor (the most abundant 5-HT receptor in the hippocampus), and the 5-HT_2A_ receptor are highly associated with GR- and MR-producing neurons in the human hippocampus, which control glucocorticoid secretion[96, 106]. As such, it could be hypothesized that acute agonism of these 5-HT receptors stimulates the HPA axis and increases cortisol, as seen in ours and the previously mentioned studies. Subsequent persisting blunting of the HPA axis[107] could then be due to agonist-induced 5-HT_2A_ and 5-HT_1A_ receptor downregulation and their interaction with glucocorticoid controlling receptors (GR and MR). All that said, there is both a psychological aspect to stress, i.e. *perceiving* something as stressful due to past experiences and current coping mechanisms, and the resulting neurobiological and neuroendocrine response[94]. If psilocybin does alter the stress response, our data do not allow us to assess at what stage psilocybin exerts an effect-the neurocognitive appraisal of the stressor or the subsequent HPA axis activity. Our data may suggest an interaction with the latter, as there were no changes on reports of trait anxiety independent from the stress test, however future studies with a larger sample size should investigate. Finally, as to whether changes in circulating cytokines relate to alterations in an individual’s appraisal and response to a psychosocial stressor, our data indicated a potential relationship (see supplementary analysis; Table S6). However such results must be taken with extreme caution, as the current study was not powered to confidently assess such a relationship. Thus future studies, preferably in a clinical population, should assess whether psychedelic-induced changes in the inflammatory profile underlie alterations in an individual’s response to stress.

## Conclusion

In conclusion, our findings demonstrate a rapid and persisting decrease in cytokine concentrations upon psilocybin administration (Figure 5). This acute change may contribute to the psychological and therapeutic effects of psilocybin demonstrated in ongoing patient trials. Such rapid effects may be modulated via an acute glutamatergic – TNF-α interaction in the hippocampus, whereas persisting changes in IL-6 and CRP may contribute to reported increases in mood and prosocial behavior.

**Figure 5.**
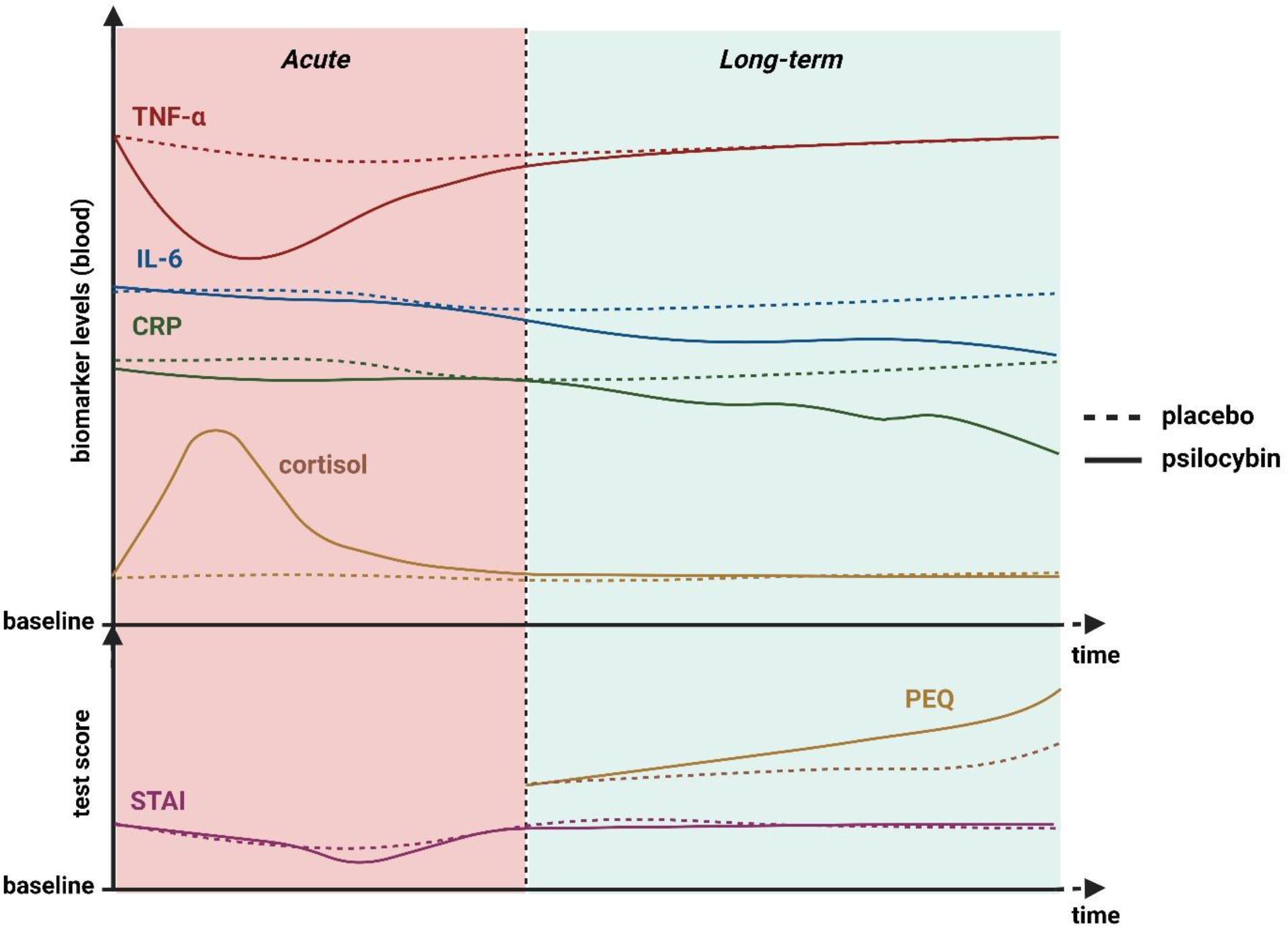
Pictorial summary of the potential connections between the biological markers assessed in this study (inflammatory and HPA-axis modulation) and the psychological outcomes (PEQ and trait anxiety). Not represented is the neuroendocrine response to the stress test, which can be found in Figure 3.

Additionally, our results suggest that even a moderate dose of psilocybin, in healthy volunteers, induces persisting alterations to HPA-axis activity in response to a psychosocial stressor. Future studies in a clinical population should confirm whether such a response relates to the symptomatic relief currently being demonstrated in clinical trials.

## Data Availability

All data produced in the present study are available upon reasonable request to the authors, after the manuscript has been published in a peer-reviewed journa;

