## supplement for "Psilocybin induces acute and persisting alterations in immune status and the stress response in healthy volunteers"

Table S1. Mean subject characteristics (SD) and history of drug use for healthy participants in the psilocybin and the placebo condition (N=60), as previously published in Mason, Kuypers [1]

| Variable | Psilocybin | Placebo | Value | *P* value |
|---|---|---|---|---|
| Sex (male/female), n, total | 18/12, 30 | 17/13, 30 | $\chi2$ =0.07 ‡ | 0.79 |
| Age, years | 22.73 (2.90) | 23.20 (3.65) | *t*=-0.55† | 0.60 |
| History of psychedelic use, years | 2.92 (2.62) | 2.19 (2.55) | *t*=1.04 | 0.30 |
| Lifetime psychedelic use, number of occasions | 9.53 (16.81) | 5.47 (8.24) | *t*=1.09 | 0.28 |
| Cannabis consumption, per month | 2.67 (3.14) | 3.24 (4.91) | *t*=-0.47† | 0.64 |
| Alcohol consumption, glasses per week | 5.47 (4.78) | 5.82 (4.02) | *t*=0.31† | 0.76 |
| Caffeine consumption, glasses per week | 10.00 (7.75) | 9.38 (8.10) | *t*=0.30† | 0.76 |
| Nicotine consumption, cigarettes per week | 3.82 (15.66) | 7.67 (15.49) | *t*=-0.96† | 0.34 |

*Significant *P* values

†Independent *t* test

‡$\chi2$ test for frequency data

Table S2. IL-6 values of one outlier participant in the psilocybin condition, compared to the group mean (SD).

| | **Variable** | **Outlier** | **Psilocybin mean (SD)** | **Placebo mean (SD)** |
|---|---|---|---|---|
| Raw value | IL-6 baseline | 12.08 | 1.30 (1.13) | 0.78 (0.41) |
| | IL-6 acute | 11.80 | 1.255 (0.76) | 0.85 (0.47) |
| | IL-6 follow-up | 3.12 | 1.06 (1.00) | 1.18 (0.87) |
| | IL-6 acute change score | -0.28 | -0.04 (0.71) | 0.07 (0.38) |
| | IL-6 follow-up change score | -8.96 | -0.21 (1.62) | 0.31 (0.73) |
| Log transformed value | IL-6 baseline | 1.08 | -0.005 (0.31) | -0.16 (0.23) |
| | IL-6 acute | 1.07 | 0.02 (0.27) | -0.13 (0.25) |
| | IL-6 follow-up | 0.49 | -0.09 (0.38) | -0.02 (0.28) |
| | IL-6 acute change score | -0.01 | 0.03 (0.24) | 0.03 (0.22) |
| | IL-6 follow-up change score | -0.59 | -0.08 (0.53) | 0.11 (0.21) |

Table S3. Mean (SE) of cytokines which did not show a treatment effect or treatment*session interaction.

| Variable | Baseline sample | | Acute sample | | Follow-up sample | |
|---|---|---|---|---|---|---|
| | Psilocybin | Placebo | Psilocybin | Placebo | Psilocybin | Placebo |
| IL-8 | 0.39 (.05) | 0.31 (.07) | 0.21 (.09) | 0.19 (.08) | 0.36 (.07) | 0.41 (.05) |
| IL-1b | 0.14 (.02) | 0.18 (.03) | 0.13 (.02) | 0.20 (.02) | 0.15 (.02) | 0.18 (.02) |

Table S4. Anxiety ratings on the POMS 50 minutes after the stress paradigm, per treatment group and per stress condition. Values are change scores from baseline.

| Treatment | Anxiety ratings (change from baseline) | | t | p | d |
|---|---|---|---|---|---|
| | Stress condition | No stress condition | | | |
| Psilocybin | -0.53 (0.33) | -0.27 (0.53) | -0.42 | 0.67 | 0.15 |
| Placebo | 1.14 (0.44) | -0.71 (0.79) | 2.05 | 0.05 | 0.77 |

Table S5. Persisting effects questionnaire at 1-week after psilocybin session. *p<0.05

| **Subscale** | **Psilocybin mean (SE)** | **Placebo mean (SE)** | ***t*-value** | **P value** | ***d*** |
|---|---|---|---|---|---|
| Positive | 19.60 (2.71) | 5.55 (1.65) | 4.42 | <.001* | 1.15 |
| Negative | 1.63 (0.50) | 0.44 (0.35) | 1.93 | 0.06 | 0.5 |
| Positive | 14.83 (2.14) | 4.00 (1.49) | 4.45 | <.001* | 1.08 |
| Negative | 1.83 (0.58) | 0.75 (0.41) | 1.50 | 0.14 | 0.39 |
| Positive | 10.30 (1.90) | 2.27 (0.78) | 3.90 | <.001* | 1.02 |
| Negative | 0.90 (0.36) | 0.24 (0.13) | 1.71 | 0.10 | 0.46 |
| Altruistic/positive | 10.53 (1.84) | 2.37 (1.10) | 3.80 | <.001* | 0.98 |
| Antisocial/negative | 1.40 (0.65) | 0.69 (0.49) | 0.87 | 0.39 | 0.23 |
| Positive | 1.87 (0.23) | 0.31 (0.14) | 5.70 | <.001* | 1.22 |
| Negative | 0.00 (0.00) | 0.07 (0.07) | -1.00 | 0.33 | Cannot compute |
| Increased | 24.40 (4.97) | 6.41 (2.91) | 3.12 | 0.003* | 0.81 |
| Decreased | 3.00 (1.22) | 0.52 (0.38) | 1.94 | 0.06 | 0.50 |

**Methods**

**Participants.** Participants were recruited through advertisements around Maastricht University and internet forums in the Netherlands. Inclusion criteria were: age, 18-40 years; previous experience with a psychedelic drug, but not within the past 3 months; normal weight, body mass index between 18 and 28 kg/m2; free from psychotropic medication; good physical health, including absence of major medical, endocrine, and neurological conditions; and written informed consent. Exclusion criteria were: history of drug abuse or addiction; pregnancy or lactation; health issues including hypertension (diastolic >90 and systolic >140), cardiac dysfunction, and liver dysfunction; current or history of psychiatric disorders; previous experience of serious side effects to psychedelics; and MRI contraindications. Before inclusion, subjects answered medical questionnaires about their health and drug use, and were screened and examined by a study physician, who checked for general health, conducted a resting ECG, and took blood and urine samples in which hematology, clinical chemistry, urine, and virology analyses were conducted. Participant demographic data can be found in Table S1, and in the previous publication[1].

Psilocybin (powder) was obtained from GH Pharm GmbH, Frankfurt, Germany. A permit for obtaining, storing, and administering psilocybin was obtained from the Dutch Drug Enforcement Administration. Participants were financially compensated for their participation in the study.

**Randomization and blinding.** An experimenter who was not responsible for treatment randomization or preparation, recruited all participants. A separate experimenter, who did not come in direct contact with the subjects, Allocated treatment in a completely random order. This latter experimenter was also responsible for preparing the treatment (psilocybin or placebo), and giving the treatment to the data collector in a closed cup. The closed cup ensured that neither the data collector or the participant would be unblinded as to what treatment the participant was receiving. Only following completion of all data collection, was the study fully unblinded.

**Procedure.** Participants were familiarized with the test day procedures on a separate training day prior to the treatment conditions. Participants were instructed to refrain from drug use, including psychedelic drugs (≥ 3 months), MDMA/ecstasy (≥ 14 days), alcohol (≥24 hours), and all other drugs of abuse (≥7 days) prior to their first testing day, and to remain sober until completion of the follow-up testing day (7 days later). Additionally, participants were asked to refrain from caffeine and nicotine use the day of the test day.

On arrival of a test day, absence of drug and alcohol use was assessed via a urine drug screen and a breath alcohol screen. An additional pregnancy test was given if participants were female. If all tests were found to be negative, participants were allowed to proceed, and a venal catheter was placed, in order to take blood samples throughout the testing day. Before administration of treatment, a baseline blood sample was taken and baseline vital signs (blood pressure and heart rate) were measured. After measurements, the treatment was administered orally, in a closed cup containing bitter lemon (placebo) or bitter lemon and psilocybin (powder; 0.17 mg/kg psilocybin). Bitter lemon was used in order to conceal any potential taste of psilocybin. After 40 minutes, participants were placed in the MRI scanner, where resting state scans and magnetic resonance spectroscopy were performed throughout a 1 hour time window. When they were taken out of the MRI scanner, they returned to the laboratory room where they completed the creativity tests. At the end of the test day (approximately 6 hours after treatment administration), participants were asked to complete measures of retrospective subjective high. Participants stayed under supervision until the testing day was complete, and the researcher deemed they were fit to go home.

Participants then returned 7 days later, and were screened again for drug and alcohol use, and if applicable pregnancy, via the aforementioned methods. If the tests were found to be negative, they were allowed to proceed. If they were positive, participants were sent home and excluded from the study (n=0).

**MRS Acquisition.**

Anatomical ($T_1$-weighted) images were acquired using magnetisation-prepared 2 rapid acquisition gradient-echo (MP2RAGE)[2] sequence (TR = 4.5 s, TE = 2.39 ms, $TI_1$ = 0.90 s, $TI_2$ = 2.75 s, flip $angle_1$ = 5°, flip $angle_2$ = 3°, voxel size = 0.9 mm isotropic, matrix size = 256 × 256 × 192, phase partial Fourier = 6/8, GRAPPA factor = 3 with 24 reference lines, bandwidth = 250 Hz/pixel, acquisition time = 6:00 min). Tissue probability maps for grey matter (GM), white matter (WM) and cerebrospinal fluid (CSF) were generated from the $T_1$-weighted anatomical images using FSL-FAST[3], and assessed for differences in spectroscopic voxels, between groups.

Single-voxel proton magnetic resonance spectroscopy (MRS) measurements were performed on a MAGNETOM 7T MR scanner (Siemens Healthineers, Erlangen, Germany) with a whole-body gradient set (SC72; maximum amplitude, 70 mT/m; maximum slew rate, 200 T/m/s) and using an single-channel transmit/32-channel receive head coil (Nova Medical, Wilmington, MA, USA). Spectroscopic voxels of interest were placed by a trained operator at the medial prefrontal cortex (voxel size = 25 mm x 20 mm x 17 mm) and the right hippocampus (voxel size = 37 mm x 15 mm x 15 mm). Spectra were acquired with stimulated echo acquisition mode (STEAM)[4] sequence using the following parameters: TE = 6.0 ms, TM = 10.0 ms, TR = 5.0 s, NA = 64, flip angle = 90°, RF bandwidth = 4.69 kHz, RF centred at 2.4 ppm, receive bandwidth = 4.0 kHz, vector size = 2048, 16-step phase cycling, acquisition time = 5:20 min. Water suppression was achieved by variable power RF pulses with optimised relaxation delays (VAPOR)[5]. In addition, a complete phase cycle of measurements was acquired without the water suppression RF pulses to record a water peak reference for eddy current correction[6] and absolute metabolite concentration calibration[7, 8]. Before the spectroscopy measurements, a 3D-GRE dual-echo field-map ($TE_1$ = 1.00 ms, $TE_2$ = 2.98 ms, TR = 20.0 ms, flip angle = 8°, voxel size = 3 mm isotropic, matrix size = 84 × 84 × 56, bandwidth = 1450 Hz/pixel, acquisition time = 2:24 min) was acquired and used to calculate the shim currents required to homogenise the static magnetic field in the spectroscopic voxels of interest.

The spectra were analysed with LCModel version 6.3-1H using a GAMMA[9] simulated basis set which includes Alanine (Ala), Ascorbic Acid (Asc), Aspartate (Asp), Creatine (Cr), γ-Aminobutyric Acid (GABA), Glucose (Glc), Glutamate (Glu), Glutamine (Gln),

Glycerophosphocholine (GPC), Glutathione (GSH), Glycine (Glyc), Lactate (Lac), Myo-Inositol (mI), N-Acetyl Aspartate (NAA), N-Acetyl Aspartyl Glutamate (NAAG), Phosphocreatine (PCr), Phosphorylcholine (PCh), Phosphorylethanolamine (PE) , Scyllo-Inositol (Scyllo), and Taurine (Tau)[10]. The metabolite basis set also includes an in vivo Macromolecules (MMol) spectrum which was collected using a metabolites suppressed double inversion recovery (DIR) STEAM with the same parameters as above and $TI_1$ = 2.09 s and $TI_2$ =0.52 s[11]. All metabolite concentrations were reported with respect to total creatine. As metabolites were reported as a ratio to tCR, absolute metabolite concentration of tCR was checked in each brain region, for significant difference between groups. There was no significant difference between groups of tCR in either the mPFC or the hippocampus suggesting that any reported significant relative metabolite concentrations were not due to an increase or decrease in tCR

**MRS Quality**. To ensure data quality and reliable metabolite estimation, only absolute metabolite values with a relative Cramer–Rao lower bound below 20%, a signal-to-noise ratio (SNR) greater than 10, and a full-width at half-maximum peak height (FWHM) < 0.1 were considered[17-19]. MRS voxel placement can be found in Supplementary Figure S2, and mean SNR, %CRLB, and FWHM values can be found in Table 1 in the original publication[1]. An overview of data points that were *not* included in the analysis, and reason why, can be found in Supplementary Table S6 in the original publication[1].

***Statistical analyses***

Further exploratory analysis were run to assess the relationship between individual cytokines, and between changes in cytokines and the response to the stress test. Spearman's correlations were conducted to assess whether there was a relationship between acute psilocybin-induced changes in TNF-α and cortisol, and persisting psilocybin-induced changes in CRP and IL-6. A canonical correlation was conducted to evaluate the association between changes in concentrations of immune functioning which showed a treatment effect (i) and changes in response to the stress test which showed a treatment effect (ii). The variables were separated into two sets; set 1 included concentrations of immune functioning (i) and set 2 included changes in cortisol and blood pressure (ii). The canonical correlation analysis was deemed as exploratory as the sample size was too low to confidently assess the relationship.

**Results**

***Acute psilocybin-induced changes in TNF-α correlated with changes in cortisol.***

There was a positive correlation between acute changes in TNF-α and acute changes in cortisol. The relationship was significant at 80 minutes post psilocybin administration, when cortisol levels were the highest ($R=.456$, $p=.02$, $n=27$), and failed to reach significance at 150 minutes ($R=.347$, $p=.08$, $n=26$).

***Persisting psilocybin- induced changes in CRP correlated with changes in IL-6.***

There was a strong positive correlation between persisting changes in CRP and IL-6 ($R=.751$, $p<.001$, $n=21$), but not between acute changes in TNF-α and persisting changes in CRP ($R=-.175$, $p=.45$, $n=21$) or IL-6 ($R=.017$, $p=.93$, $n=26$).

***Psilocybin induced changes in inflammatory markers predict physiological responses on the stress test.***

A canonical correlation analysis was conducted using the three markers of immune functioning (change scores from baseline) as predictors of the neuroendocrine stress response. The analysis yielded three functions with squared canonical correlations ($R_c^2$) of .805, .574, and .385 for each successive function. The full model across all functions was statistically significant $F(9, 12.32) = 3.28$, p=.03, explaining 94.91% of the variance. From this model, all functions were considered noteworthy, explaining 80.5%, 57.4%, and 38.5% of the variance, respectively.

Table S5 presents the standardized canonical function coefficients, the structure coefficients ($r_s$), and the squared structure coefficients ($r_s^2$) for functions 1 and 2, as well as the communalities ($h^2$) across the two functions for each variable. Function 1 indicated that the dominant contributor was the systolic blood pressure change in response to the stress test, and the secondary contributor was the diastolic blood pressure change, whereas the dominant predictor was acute changes in TNF-alpha. Overall this function suggests that the more of a change in diastolic and systolic blood pressure in response to the stress test, the lower the levels of TNF-alpha.

Function 2 indicated that the dominant contributor was changes in diastolic blood pressure and the secondary contributor were changes in cortisol in response to the stress test, whereas the dominant predictor was persisting changes in IL-6 concentration levels, and the secondary predictor were acute changes in TNF-alpha. Overall this function suggests that the higher the diastolic blood pressure change in response to the stress test, the lower the levels of IL-6 and TNF-alpha.

Function 3 indicated that the dominant contributor was peak changes in cortisol concentrations in response to the stress test, whereas the dominant predictors were persisting changes in CRP and IL-6. Overall this function suggests that the more of a change in cortisol, the more of a reduction of IL-6 and CRP.

Table S6. Canonical solution for inflammatory markers predicting response to the stress test.

| Variable | Function 1 | | | Function 2 | | | Function3 | | | |
|---|---|---|---|---|---|---|---|---|---|---|
| | Coef | $r_s$ | $r_s^2$ (%) | Coef | $r_s$ | $r_s^2$ (%) | Coef | $r_s$ | $r_s^2$ (%) | $h^2$(%) |
| Peak cortisol change | .291 | -.057 | 0.32 | .403 | .463 | 21.43 | .938 | .884 | 78.14 | 99.89 |
| Diastolic change | -.204 | .551 | 30.36 | -1.226 | -.814 | 66.25 | .629 | .179 | 3.20 | 99.81 |
| Systolic change | 1.18 | .949 | 90.06 | .736 | -.253 | 6.40 | -.308 | -.185 | 3.42 | 99.89 |
| $R_c^2$ | | | 80.51 | | | 57.49 | | | 38.56 | |
| TNF alpha acute change | -1.135 | -.772 | 59.59 | .071 | .540 | 26.16 | .254 | .333 | 11.08 | 96.83 |
| IL-6 longterm change | 1.73 | -.108 | 1.08 | 2.15 | .717 | 51.41 | .514 | -.687 | 47.19 | 99.68 |
| CRP longterm change | -1.71 | -.181 | 3.27 | -1.59 | .364 | 13.25 | -1.38 | -.913 | 83.35 | 99.87 |
